# The association between maternal intra-abdominal pressure, pregnancy induced hypertension

**DOI:** 10.1101/2023.03.28.23287874

**Authors:** Sajith Jayasundara, Malik Goonewardene, Lanka Dassanayake

## Abstract

**Introduction:** Pregnancy leads to a state of chronically increased intra-abdominal pressure (IAP) caused by a growing fetus, fluid, and tissue. Increased intra-abdominal pressure is leading to state of Intra-Abdominal Hypertension (IAH) and Abdominal Compartment Syndrome. Clinical features and risk factors of preeclampsia is comparable to abdominal compartment syndrome. Intra-abdominal pressure (IAP) may be associated with the pathogenesis of pregnancy induced hypertension.

**Objectives:** The study aimed to determine the antepartum and postpartum IAP levels in women undergoing caesarean delivery (CD) and association between hypertension in pregnancy, and antepartum and postpartum IAP levels in women undergoing caesarean delivery (CD).

**Method:** Seventy pregnant women (55 normotensive, 15 hypertension in pregnancy) undergoing antepartum, non-emergency CD, had their intravesical pressure measured before and after the CD, the intravesical pressure measurements obtained with the patient in the supine position were considered to correspond to the IAP. Multivariable linear regression models were used to study associations between intraabdominal pressure and baseline characteristics in normotensive pregnancies and hypertensive pregnancies.

**Results:** In normotensive pregnancies at mean gestation age of 38 weeks +2days (95%CI 37+6 to 38 +4), mean antepartum IAP was 12.7 mmHg(95%CI 11.6 to 13.8) and the mean postpartum IAP was 7.3 mmHg (95% CI 11.6 to 13.8).

Multivariable linear regression models showed hypertension in pregnancy group antepartum IAP positively associated with coefficient value of 1.617 (p= 0.268) comparing with normotensive pregnancy group.

Postpartum IAP in hypertension in pregnancy group positively associated with coefficient value of 2.519 (p= 0.018) comparing with normotensive pregnancy group.

IAP difference is negatively associated with hypertension in pregnancy (coefficient -1.013, p= 0.179)

**Conclusion:** In normotensive pregnancies at term, the IAP was in the IAH range of the non-pregnant population. Higher Antepartum IAP and Postpartum IAP are associated with hypertension in pregnancy. Reduction of IAP from antepartum period to postpartum period was less with hypertension in pregnancy.

## Introduction

Pregnancy with growing fetus and amniotic fluid, leading to increased pressure within the abdominal cavity. The importance of the diagnosis and management of intra-abdominal hypertension (IAH) and abdominal compartment syndrome is increasingly recognized in different fields of medicine. To date, little is known about normal values of intra-abdominal pressure (IAP) during pregnancy either in healthy or complicated pregnancies.

Despite decades of research into the etiology and mechanism of preeclampsia, its exact pathogenesis remains uncertain. Several authors have hypothesized that Intra-abdominal hypertension (IAP) as an etiologic factor of pregnancy induced hypertension (PIH). Risk factors and clinical manifestation of abdominal compartment syndrome are similar to the preeclampsia and eclampsia. Supporting this hypothesis is the fact that PIH is more often seen in association with a first pregnancy (when the abdomen has not been previously stretched) than subsequent pregnancies, with twin pregnancies which would be more likely to be associated with an increased IAP and with obesity [1]. An animal study has shown that chronically increased intra-abdominal pressure would lead to systemic hypertension [2].

In the 1900s, Paramore had suggested uncompensated elevated IAP as a possible etiologic factor in the development of preeclampsia [3].

Harvey Sugerman hypothesized that preeclampsia is a venous disease secondary to an increased intra-abdominal pressure. Pregnancy with high IAP may produce all of the pathology associated with preeclampsia secondary to compression, producing decreased flow in the venous system throughout the body [4]. It is postulated that this decreased venous flow increases progressively throughout the body as the IAP rises, leading to lower body edema, fetal ischemia and placental ischemia, pre-mature maturation and infarction, decreased renal venous flow leading to activation of the juxtaglomerular apparatus, hypertension and proteinuria, decreased portal and splenic venous flow with hepatic ischemia, necrosis, elevated liver enzymes (transaminases), hypersplenism with thrombocytopenia and hemolysis, increased intra-thoracic pressure with upper body edema, decreased jugular venous flow, increased intra-cranial pressure, and seizures.

## Method

Ethical approval was obtained from Ethical Review Committee, Faculty of Medicine, University of Ruhuna, Galle, Sri Lanka. Informed written consent was obtained from all the participants in the study. The enrolment was carried out in 2016 and 2017.

A cross sectional study was carried out on a cohort of 70 women with singleton pregnancies, undergoing antepartum, non-emergency Caesarean Delivery (CD) in the Academic Obstetric Unit, Teaching Hospital, Mahamodara, Galle, Sri Lanka. Pregnant women underwent Category 1 caesarean delivery and caesarean delivery performed during active phase of labour, are excluded.

Antenatal care data was collected to data sheet, from in-ward clinical records, antenatal care records and investigation results.

The participants were transferred to the operating room and the pre-operative blood pressure was recorded. Prior to spinal or general anaesthesia, patients were placed in the supine position, and a transurethral 16-Fr Foley catheter was inserted into the bladder with non-touch technique under aseptic conditions. Urinary bladder was emptied and the sample of urine checked for albumin. Intra vesical pressure measurements was taken using the Intravesical pressure measurement system according to the world abdominal compartment society recommendation with patient in supine position, mid axillary level as zero reference and 25ml of bladder inflation volume.

All the measurements were obtained by the principal investigator. Three consecutive measurements were obtained at the end of expiration and mean value of the three measurement was calculated. Measurements was repeated between six to eight hours post-operatively. Birth weight of the babies were measured using same standard weight measuring scale.

The Statistical Package for Social Sciences (SPSS 22.0; SPSS Inc., Chicago, IL) version 22.0 was used for statistical analysis. Data normality was assessed by the Kolmogorov-Smirnov test. Statistical comparisons among groups were performed using the independent *t* test and Chi-square test. All *p* values were two tailed and statistical significance was set as p<0.05.

The effect of belonging in the hypertensive group on the dependent variables antepartum IAP, postpartum IAP and difference between antepartum IAP and postpartum IAP was estimated using least squares multivariable regression analyses, adjusting for maternal BMI, birthweight, gestational age and parity. Linear regression analysis was used as the dependent variable is continuous in all three cases (antepartum IAP, postpartum IAP, IAP difference). Firstly, a pearson correlation analysis was conducted in order to test for pairwise correlations between BMI, birthweight and gestational age. Birthweight and gestational age were found to be highly positively correlated (correlation coefficient: 0.78) therefore it was decided that birthweight would need to be retained and gestational age would need to be removed from all subsequent regression analyses.

For each of the three dependent variables, different combinations of regression models were fit in order to assess all possible combinations of variables and choose the model with the highest adjusted R^2^ value. The adjusted R^2^ value is a robust goodness-of-fit metric in linear regression models and shows the percentage of the variance of the dependent variable that is attributed to the independent variables, accounting for the number of variables in the model. Squared transformations of variables in the regression models were also examined in order to check for improvements in model fitting. Hypertension was included in the models as a binary categorical variable; hypertensive subjects were assigned the value 1 while normotensive the value 0. A p value lower than 0.05 corresponding to each coefficient in the model was used as a criterion for significance.

## Results

Seventy subjects who undergoing caesarean delivery (CD) were enrolled. Subjects were categorized into, normotensive (n=55, 78.6%) and Hypertension in pregnancy (HIP) (n= 15, 21.4%). HIP include Gestational hypertension (n=5), Preeclampsia (n= 8), Eclampsia (n=1) and chronic hypertension (n=1). Mean values with standard deviations and 95% confidence intervals or Median with interquartile range calculated in each parameter calculated in each category as shown in Table 1.

**Table 1.** Characteristics of subjects.

|  | Normotensive<br>(n=55) | Hypertension in Pregnancy<br>(n=15) | *p* value |
| --- | --- | --- | --- |
| Age ( Years) |  |  |  |
| Mean( 95% CI) | 32.5 (31.4 to 33.7) | 32.9 (29.8 to 35.9) | 0.805* |
| GA (weeks + days) |  |  |  |
| Mean (95% CI) | 38+1(37+6 to 38+4) | 34+2 (32+3 to 36+1) | 0.000* |
| Parity |  |  |  |
| Median( IQR) | 2.0(1) | 2.0 (3.0) | 0.220** |
| BMI ( kg/m<sup>2</sup> ) |  |  |  |
| Mean (95% CI) | 23.6 (22.9 to 24.3) | 24.5 (23.1 to 25.9) | 0.241* |

CI = Confidence interval GA = Gestational age IQR = Interquartile Range \* Independent t test
\*\* Chi-square test

Outcome measures in normotensive category and hypertension in pregnancy category compared with Systemic blood pressures, IAP parameters, and birth weight of the neonates. Mean values with 95% Confidence interval calculated in each parameter calculated in each category as shown in Table 2.

**Table 2.** Systematic blood pressure and Intra-abdominal pressure in normotensive category and hypertension in pregnancy category.

|  | Normotensive<br>(n=55) | Hypertension in<br>Pregnancy (n=15) | *p* value |
| --- | --- | --- | --- |
| Birth weight (g) |  |  |  |
| Mean | 2934.5 | 1886.7 | 0.000* |
| 95% CI | 2798.6 to 3070.5 | 1415.9 to 2357.4 |  |
| Antepartum MAP (mmHg) |  |  |  |
| Mean | 87.5 | 122.2 | 0.000* |
| 95% CI | 85.9 to 89.1 | 114.5 to 129.9 |  |
| Postpartum MAP (mmHg) |  |  |  |
| Mean | 86.0 | 101.3 | 0.000* |
| 95% CI | 84.4 to 87.7 | 94.4 to 108.2 |  |
| Antepartum IAP (mmHg) |  |  |  |
| Mean | 12.7 | 12.5 | 0.892* |
| 95% CI | 11.6 to 13.8 | 10.0 to 15.0 |  |
| Postpartum IAP (mmHg) |  |  |  |
| Mean | 7.3 | 9.2 | 0.022* |
| 95% CI | 6.5 to 8.0 | 7.3 to 11.2 |  |
| IAP difference<sup>a</sup> (mmHg) |  |  |  |
| Mean | 5.4 | 3.3 | 0.002* |
| 95% CI | 4.8 to 6.0 | 2.1 to 4.6 |  |

95%CI= 95% Confidence interval MAP=Mean arterial pressure, IAP=Intra-abdominal pressure a - IAP difference = Pre caesarean Delivery intra-abdominal pressure – Post caesarean delivery intra-abdominal pressure \* Independent t test

Different combinations of variables in each model were tested in order to retain the most efficient model for each dependent variable. The most robust and parsimonious regression model, i.e. the model with the highest adjusted R^2^ (0.032), when considering the effect of the aforementioned independent variables on antepartum IAP included hypertension and birthweight as independent variables. Birthweight (p = 0.042) was found to be a significant predictor of antepartum IAP while hypertension was found to be positively associated to antepartum IAP (coef: 1.617) although a statistically significant effect was not established (p = 0.268). Distribution of antepartum IAP level against the birth weight shown in Fig 1.

**Fig 1.**
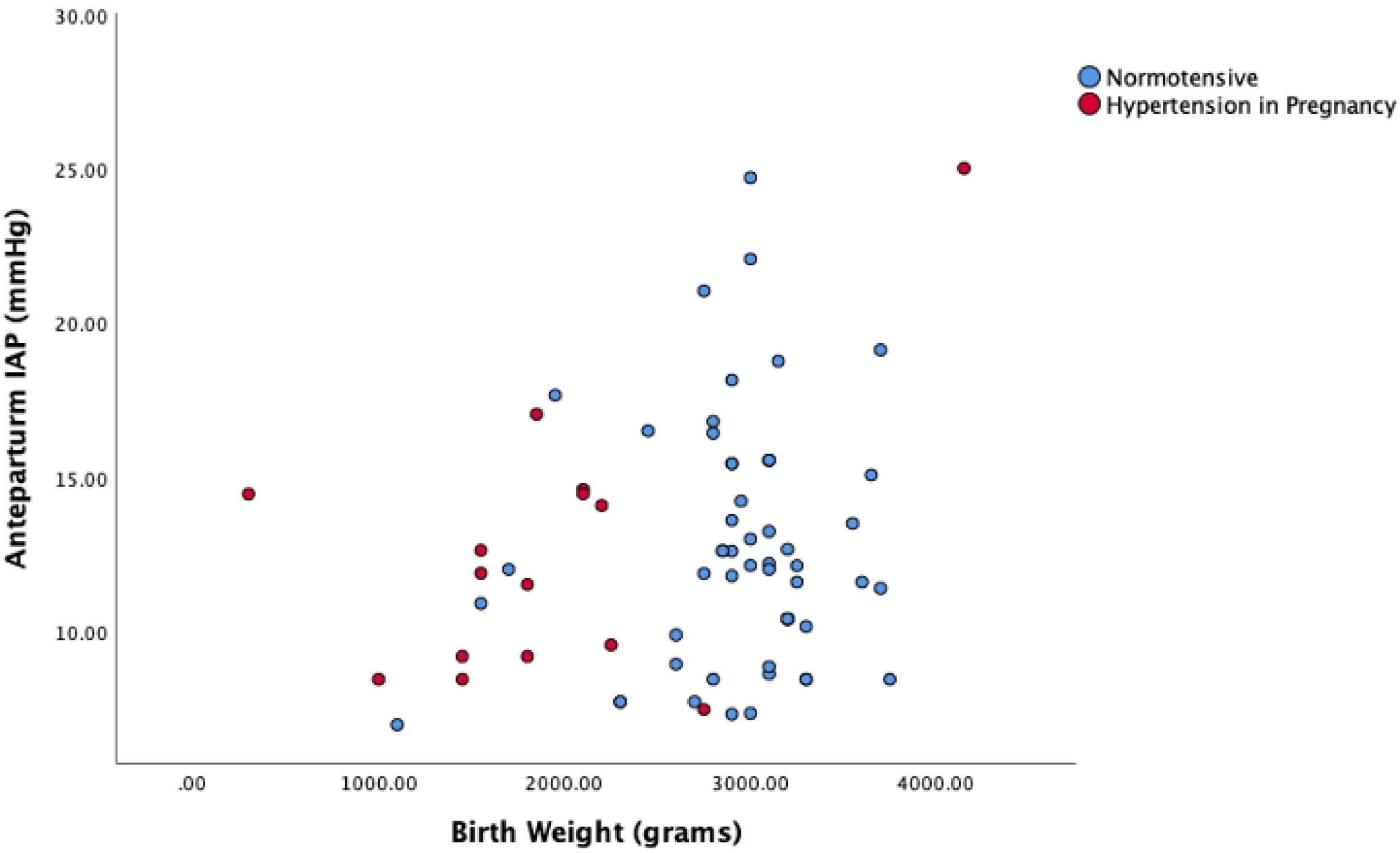
Antepartum IAP level and the birth weight in normotensive and hypertension in pregnancy. The most efficient model with respect to predicting IAP difference (R^2^: 0.190) included the variables hypertension (p= 0.179) and birthweight squared (p =0.01). Hypertension was negatively associated (coef: -1.01) to IAP difference although a statistically significant effect was not established (p = 0.179). Distribution of IAP difference level against the birth weight shown in Fig 2.

**Fig 2.**
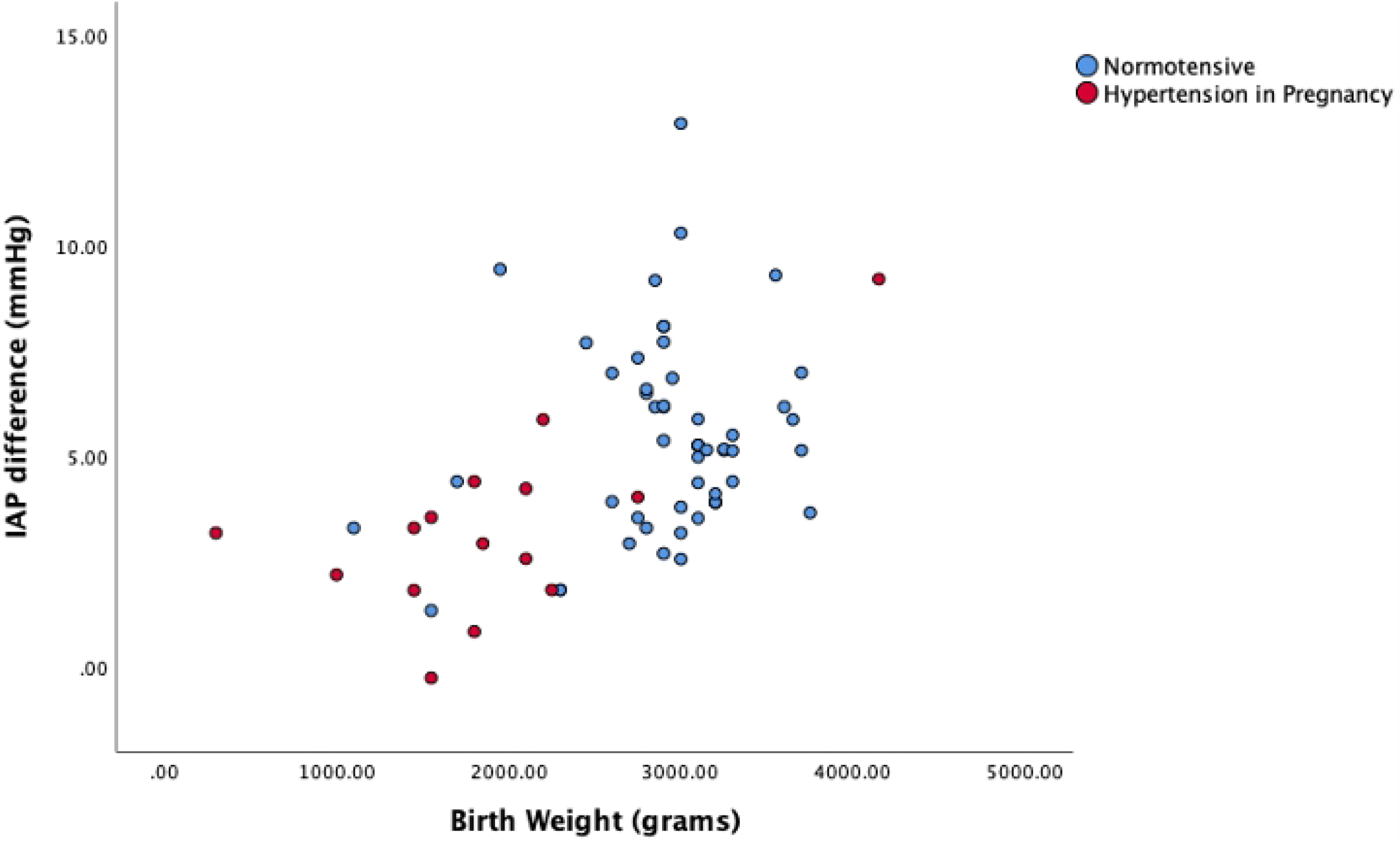
IAP difference and the birth weight in normotensive and hypertension in pregnancy. The most efficient model with respect to predicting post-IAP (R^2^: 0.059) included hypertension and birthweight. Hypertension was found to be positively (coef: 2.519) and significantly associated to postpartum IAP (p = 0.018). Distribution of postpartum IAP level against the birth weight shown in Fig 3.

**Fig 3.**
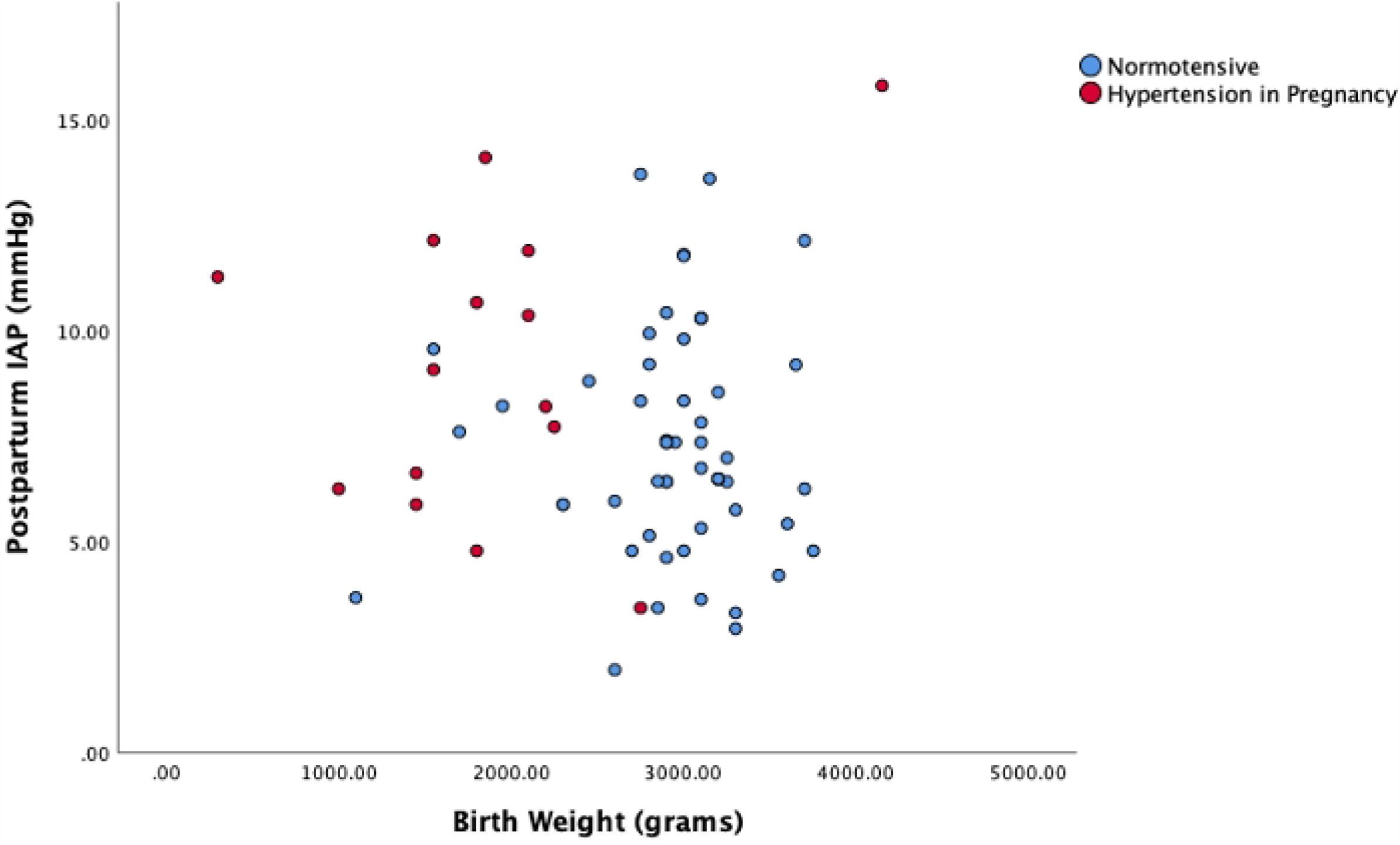
Postpartum IAP and the birth weight in normotensive and hypertension in pregnancy. Overall, for all three dependent variables, hypertension and birthweight produced the best adjusted R^**2**^ scores so these were the variables that were chosen to be included in the models.

## Discussion

### Intra-abdominal pressure in normotensive pregnancy

In this study, 55 of normotensive subjects at term, antepartum IAP was 12.69 mmHg (95% CI =11.62 to 13.77) and postpartum IAP 7.26 mmHg (6.54 to 7.98). Comparing with other five studies published regarding antepartum and postpartum IAP is shown in Table 3.

**Table 3.** **Comparison of studies of intra-abdominal pressure in normotensive pregnancies**

| Author | Our study | Fuchs F *et al*<br>2013 [5] | Chun R *et al*<br>2011 [6] | Staelens A *et al*<br>2014 [7] | Al Khan A *et al*<br>2011 [8] | Abdel-Razeq<br>*et al*<br>2010 [9] |
| --- | --- | --- | --- | --- | --- | --- |
| Country | Sri Lanka | France | Canada | Belgium | USA | USA |
| Sample size | 55 | 70 | 20 | 23 | 100 | 21 |
| Age | 32.53(31.4 to 33.7) | 34 (27-41) | 31(SD 5) | 29.3 (SD 3.5) | 31.7 (5.1) | - |
| Gestational Age (weeks) | 38.25 ( 37.9 to 38.6) | 38.1 (38 to 39) | 38.6 (SD 1.6) | 39.0 (0.6) | 38.4 ( 0.8) | - |
| Birth weight | 2934.55 ( 2799 to 3071) | 3210 (2445 to 4215) | - | 3590 (SD- 472.2) | 3422.8 ( 450.1) | - |
| **Antepartum IAP** | **12.69 ( 11.6 to 13.8)** | **14.2 (6.3 to 23)** | **10.9 ( 6.23 to 15.57)** | **14.0 (11.4 to 16.6)** | **22.0 (2.9)** | - |
| **Postpartum IAP** | **7.26 ( 6.54 to 7.98)** | **11.5 (5 to 19.7)** | - | **9.8 (6.8 to 12.8)** | **16.4 ( 2.6)** | **6.4 (1.2 to 11.6)** |
| Measuring technique | Supine, 25ml, level of mid axillary line | Supine, 25ml, level of symphysis pubis | Supine, 25ml, level of mid axillary line | Supine, 20ml, , level of mid axillary line | Supine with leftward tilt, 50ml | Supine, 25ml, symphysis pubis |

IAP – Intra abdominal pressure, SD – Standard deviation

In the current study, normotensive pregnancies at term, antepartum IAP of 12.6mmHg (95% CI =11.6-13.8) is within the range of three studies, Fuchs F et al [5],Chun R et al[6] and Staelens A et al[7]. In our study, antepartum mean IAP slightly lower than Fuchs F et al and Staelens A et al mean antepartum IAP which may attribute with birth weight and other demographic factors. Al Khan A *et al* [8] study IAP value is higher than our antepartum IAP. This higher value of IAP, probably due to methodological overestimation, as is noticed by Chun et al [6] and Staelens A. et al[7].

Postpartum IAP in this study was within the range of Postpartum IAP of Fuchs F et al [5], Staelens A et al [6], and Abdel-Razeq et al [9]studies. Al Khan A et al [8] study IAP value is higher than our postpartum IAP.

### Intra-abdominal pressure in hypertension in pregnancy

In our study, antepartum, postpartum IAP levels are 12.5 mmHg (10.04 to 15.01) and 9.21 mmHg (7.26 to 11.16) respectively at mean gestational age and 34.3 weeks and mean birth weight of 1886.66g. Antepartum IAP is within in the range of Grade 1 IAH. Postpartum IAP level slightly higher than the normal level of IAP in non-pregnant population.

Mesut A. Ünsal et al published a study comparing the IAP in hypertension in pregnancy and normotensive pregnancy [10].

Comparing with Mesut A. Ünsal et al study and this study, both Antepartum IAP and Postpartum IAP of hypertension in pregnancy category, within same range as shown in Table 4.

**Table 4.** Comparison of intra-abdominal pressure in hypertension in pregnancy.

|  | Mesut A. Ünsal et al<br>2016 | Current study |  |
| --- | --- | --- | --- |
| Category | Hypertension in<br>pregnancy<br>n=35 | Hypertension in pregnancy<br>n=15 | *p* value |
| Age ( 95% CI ) | 28.6 (22.4 to 34.8) | 32.87 (29.54 to 36.19) | 0.03* |
| Parity | 1 (0 to 6) | 2 |  |
| Gestational age ( 95% CI ) | 33 (24 to 40) | 34.26 (32.24 to 36.28) | 0.32* |
| BMI ( 95% CI ) | 27.0 (19.1 to 34.9) | 24.50 (23.00 to 26.00) | 0.24* |
| Birth weight ( 95% CI ) | 1910.9 (997.8 to 2824) | 1886.66 (1415.89 to 2357.44) | 0.93* |
| Ante-partum IAP -mmHg ( 95% CI ) | 13.46 (11.55 to 15.37) | 12.53 (10.04 to 15.01) | 0.30* |
| Post-partum IAP mmHg ( 95% CI ) | 10.00 (7.57 to 12.43) | 9.21 (7.26 to 11.16) | 0.36* |

IAP- Intra abdominal pressure BMI – Body Mass Index \* Independent *t* test

Multivariable regression analyses were used to compare the IAP levels in two categories. Because two groups have different birth weights and gestational age which directly influence the IAP levels.

Regression analysis of antepartum IAP showed 1.6 times higher IAP level in hypertension in pregnancy compared to the normotensive group when adjusted for birthweight. However, it is not statistically significant, which is probably due to the small sample size in the hypertensive group. Association between IAP and hypertension in pregnancy, can be better explained with serial estimation of antepartum IAP in different gestations using a larger sample.

Reduction of IAP from antepartum period to postpartum period is 1.013 times less in hypertension in pregnancy, compared to normotensive pregnancy when adjusted for birthweight.

Postpartum IAP level is 2.5 times higher in hypertensive pregnancies compared to normotensive pregnancies after adjusting for birth weight, BMI and parity. It is statistically significant.

Delivery of the fetus and placenta is considered as the curative treatment in preeclampsia. This is based on theory of “preeclampsia is disease of placental origin”. This theory is challenged in the situation of postpartum preeclampsia, eclampsia and de novo postpartum hypertension. It raises the possibility of a residual factor or pathological process that exist absence of the fetus and the placenta. Our study revealed higher postpartum IAP level and less reduction of IAP level from antepartum period to postpartum period in hypertension in pregnancy. It infer the association of high postpartum intra-abdominal pressure and development of postpartum preeclampsia, eclampsia and de novo postpartum hypertension.

## Conclusion

In normotensive pregnancies at term, mean antepartum IAP was 12.69mmHg (95%CI 11.61 to 13.77) and postpartum mean IAP was 7.26mmHg (95% CI 11.61 to 13.77). In normotensive pregnancy at term, IAP is within the range of IAH of non-pregnant population.

Higher antepartum and postpartum IAP level, less reduction of IAP after delivery of the fetus and placenta is associated with hypertension in pregnancy compare to the normotensive pregnancy.

## Data Availability

All relevant data are within the manuscript.

## Acknowledgment

We would like to thank all volunteers for participation in this study. We thank Kosala Gayan Weerakoon for helpful discussions and support, and Kyriakos Kouvelakis for assistance with statistical analysis.

